# A Holistic Approach to Cognitive Frailty in Community-dwelling Older Adults in India: Evidence from a Nationally Representative Survey

**DOI:** 10.1101/2024.03.19.24304523

**Authors:** Sayani Das, Emma Pooley, Carol Holland

## Abstract

**Background:** Cognitive frailty (CF), a complex intersection of physical frailty (PF) and cognitive impairment (CI), remains inadequately explored in the Indian context. Given its potential link to dementia and reversible characteristics, this study aims to understand the national-level prevalence of CF, its regional variations, and its holistic association with socio-demographic, physical, psychosocial, and lifestyle factors. **Methods:** The present study employed data from the Longitudinal Ageing Study in India (LASI-Wave-I), a nationally representative survey that included 27,379 individuals (13,293 men and 14,086 women) aged 60+. CF was defined as the presence of both PF and CI without dementia. PF was assessed using the modified Community-Oriented Frailty Index, comprising a 20-item deficits frailty index. CI was evaluated using the composite score, which considered five cognitive domains and seven difficulties with IADL. To investigate the association between the outcome variable (CF) and predictor variables (socio-demographic, physical, psychosocial, and lifestyle factors), the study utilised stepwise binary logistic regression (backward elimination) and presented the results as adjusted odds ratios (AOR) with corresponding 95% confidence intervals. **Results:** The overall prevalence of CF amongst those aged 60+ was found to be 4.5%, with higher rates evident in the Western (6.2%) and Southern (5.3%) regions. Several factors exhibited significant associations with CF (p < 0.01), including being aged 80 and above (AOR 4.7, 95% CI: 3.9–5.6), being female (AOR 1.6, 95% CI: 1.4–1.9), and having less than primary education (AOR 15.3, 95% CI: 7.2–32.5, p < 0.01). Mobility impairment (AOR 6.7, 95% CI: 4.7–9.8) also exhibited a strong effect on CF. Psychosocial factors, such as low social participation (AOR 2.1, 95% CI: 1.8–2.4), major depression (AOR 1.7, 95% CI: 1.4–2.0), sleep problems (AOR 1.6, 95% CI: 1.4–1.9), and low life satisfaction (AOR 1.4, 95% CI: 1.2–1.6), were also found to have significant associations with CF. **Conclusions:** This pioneering study outlines the prevalence of CF in India, revealing elevated rates among the community-dwelling population aged over 60 compared to global figures, accompanied by notable regional variations, and underscores the importance of future longitudinal data and targeted investigations for informing policy implications.

## Background

India, the world’s most populous country, is currently undergoing a profound demographic transformation marked by a rapid increase in its ageing population. The nation is witnessing a significant surge in the number of older adults, typically classified as individuals aged 60 and above. According to the latest United Nations Population Fund’s (UNFPA) ageing report, India’s older population is expanding at an unprecedented rate, with 10.5% of the country’s total population falling within this demographic group [1]. Moreover, projections indicate that by 2050, India’s population will include approximately 347 million older adults, comprising more than 20.8% of the total population, signifying that one in every five individuals in India will be an older adult [1, 2]. These statistics underscore the pressing need to comprehend and address the multifaceted challenges associated with the health and well-being of this burgeoning demographic. As India’s ageing population continues to grow, it becomes increasingly critical to concentrate on holistic healthcare, early intervention, and support systems to enhance the well-being of older adults. Holistic healthcare represents an approach to wellness that goes beyond just the physical aspects of health. It encompasses a comprehensive understanding of health that addresses the interconnectedness of various dimensions, including the physical, mental, emotional, social, and spiritual aspects [3].

Cognitive impairment (CI), a condition encompassing various degrees of decline in cognitive functions such as memory, reasoning, and decision-making, is emerging as a significant health concern in India [4, 5]. The challenge is becoming increasingly prevalent given the increasing older population [6, 7, 8], necessitating a comprehensive study to understand the scope and implications of CI within the Indian context. In 2023, a national-level study highlighted that 7.14% of older men and 20.03% of older women reported CI [8]. However, the prevalence of CI varies from 3.5% to 11.5% from Northern to Southern parts of the country [7]. As India’s population ages, this prevalence is expected to increase, making CI a significant public health concern. On the other hand, frailty is also a complex and multifaceted condition that has gained increasing attention in the field of geriatric health worldwide. It is characterised by a state of vulnerability, where individuals are at an increased risk of adverse health outcomes due to a decline in physiological reserve across multiple organ systems [9, 10, 11]. Frailty is not limited to any specific age group, but it is more prevalent and pertinent in older adults [12, 13]. It has emerged as a growing concern in India, especially considering the ageing population as outlined. Drawing from a nationally representative sample of Indian adults aged 45 and above, research has estimated a 30.0% prevalence of frailty when utilising an accumulation of deficits-based frailty index comprising 40 deficits [13]. On the other hand, Das and Prasad (2023) evaluated frailty using the physical phenotype scale and observed an overall national prevalence of physical frailty: 27.9% for men and 33.2% for women among adults aged 60 years and above [14]. A global systematic review, which compared and pooled data on the prevalence of frailty among community-dwelling older people, highlighted a wide variation in frailty prevalence within the community, ranging from 4.0% to 59.1% [15]. The study concluded that the overall weighted prevalence of frailty was 10.7%, which emphasises that the prevalence of frailty in India exceeds the global average.

Cognitive frailty (CF) is a relatively recent concept in the field of geriatric medicine and public health, presenting a multifaceted condition that amalgamates elements of physical frailty and cognitive impairment, which commonly co-occur [16, 17]. While traditional frailty primarily concentrates on physical aspects, CF encompasses cognitive functions, including memory, executive function, and attention. It can be defined as a condition where individuals experience both physical frailty and cognitive impairment in the absence of dementia [16, 18]. Worldwide, sociodemographic characteristics such as age, gender, residence status, geographical region and living arrangements have been extensively studied, revealing their significant association with CF [19–24]. Higher education, occupational status, and economic standing also play pivotal roles, underscoring the complex relationships with CF [25–27], as they are associated with enhanced overall health outcomes, reflecting the influence of intellectual stimulation and access to resources. However, evidence also recognises the profound impact of environment and lifestyle choices in terms of health-related behaviour such as weight management, exercise and smoking. These choices, as well as environmentally related stresses, significantly influence vascular health, oxidative stress, and inflammation, contributing to physical frailty and variability in cognitive function. Therefore, smoking habits, nutritional patterns, and mobility intricately contribute to the link between cognitive and physical health and so CF [20, 28–30]. Life events such as bereavements or hospital stays can also have an impact on CF [31], introducing both physical and mental stressors than can affect overall health and psychosocial factors, such as depression, social participation, and sleeping habits, also have complex impacts on CF mechanisms [32–34].

This background illustrates the range of factors that influence cognitive function and overall health, and underlying mechanisms, emphasising the necessity for a comprehensive, holistic understanding. While the sharing of several risk factors for dementia [35] suggests a greater likelihood of the development of neurodegenerative conditions, it is important to note that not all individuals with CF progress to dementia. Although the CF state is significant because it elevates the risk of dementia, it is highly important because of its characterisation as a reversible state [21, 36, 37]. This quality makes CF a compelling focus for interventions aimed at mitigating its impact, suggesting that targeted strategies could be employed to address and potentially reverse CF. Understanding and addressing the factors contributing to CF could pave the way for effective interventions that enhance cognitive and physical well-being and potentially reduce the risk of dementia and later life dependence. Therefore, CF emerges as a critical area for research and intervention, emphasising the importance of early identification and management to potentially reduce the risk of dementia and enhance the quality of life for older adults [24, 38, 39]. A comprehensive understanding of CF is imperative for healthcare professionals, as it demands a multidisciplinary approach, including assessments covering both physical and cognitive domains, as well as broader psychosocial risk factors.

CF in India remains largely unexplored, lacking the depth of understanding that could be derived from a comprehensive national-level study. As a heterogeneous country with diverse geographical regions, India has already seen exploration into variations in physical frailty [13] and cognitive impairment [7]. However, the specific variation in terms of cognitive frailty has yet to be thoroughly examined. These gaps in research underscore the pressing need for increased awareness among academia, policymakers, and the broader community. To date, only a solitary cross-sectional study [23], conducted in the state of West Bengal, one of the most vulnerable states in terms of frailty in the nation [13], has ventured into this domain. This pioneering study revealed an overall prevalence of CF at 21.8% of those aged 60 years and above, illuminating the significance of this issue [23]. The study unveiled several key associations with CF status. Increasing age, gender (with women being more susceptible), non-married status, lower educational attainment, and non-working sociodemographic status were found to be significant factors linked CF. Moreover, the study also unveiled a range of challenges faced by cognitively frail participants, including poor nutritional status, diminished health-related quality of life, and a higher incidence of depression. These findings underscore the multifaceted nature of CF and emphasise the necessity for further research, awareness, and targeted interventions to address this underexplored aspect of health in India. Therefore, the present study aims to understand the national prevalence of cognitive frailty, regional variations and its holistic association with sociodemographic, physical, and psychosocial health and lifestyle factors by using a national survey in India.

## Methods

### Study sample and data selection

The study utilised data from the Longitudinal Ageing Study in India (LASI-Wave-I), a nationally representative survey encompassing more than 73,000 older adults aged 45 and above, along with their spouses (irrespective of age), across all 28 states and 8 union territories of India. Employing a multistage stratified area probability cluster sampling design, the survey aimed to ensure a diverse representation of respondents. In rural areas, a three-stage sampling design was implemented, whereas urban areas followed a four-stage sampling design. The first stage involved the selection of Primary Sampling Units (PSUs), defined as sub-districts (Tehsils/Taluks) within each state and union territory. The second stage focused on the selection of villages in rural areas and wards in urban areas within the chosen PSUs. In rural areas, the third stage involved the selection of households from the selected villages, while in urban areas, an additional stage was introduced, wherein one Census Enumeration Block (CEB) was randomly selected in each urban area. Subsequently, households were chosen from these CEBs in the fourth stage. Each consenting respondent in the selected households underwent an individual survey schedule. LASI stands as India’s premier ageing study, boasting the world’s largest sample size. Its primary goal is to furnish a comprehensive longitudinal evidence base on the health, social, and economic well-being of the older adults in India. This information serves as a foundation for formulating policies and programs directed at enhancing the overall well-being of the older population. LASI is designed to be conducted every three years for the next quarter-century, offering a unique opportunity to assess the impact of evolving policies on behavioural outcomes in India. For this study the sample of older adults aged 60 years and above were considered (N = 27,379; men, n= 13,293 and women, n = 14,086). **Figure 1** illustrates the identification of the sample, including the exclusion of participants with incomplete data. The detailed methodology of the survey, including information on survey design, data collection, and quality control measures, is available in https://lasi-india.org as well as in the published survey report [40].

**Figure 1.**
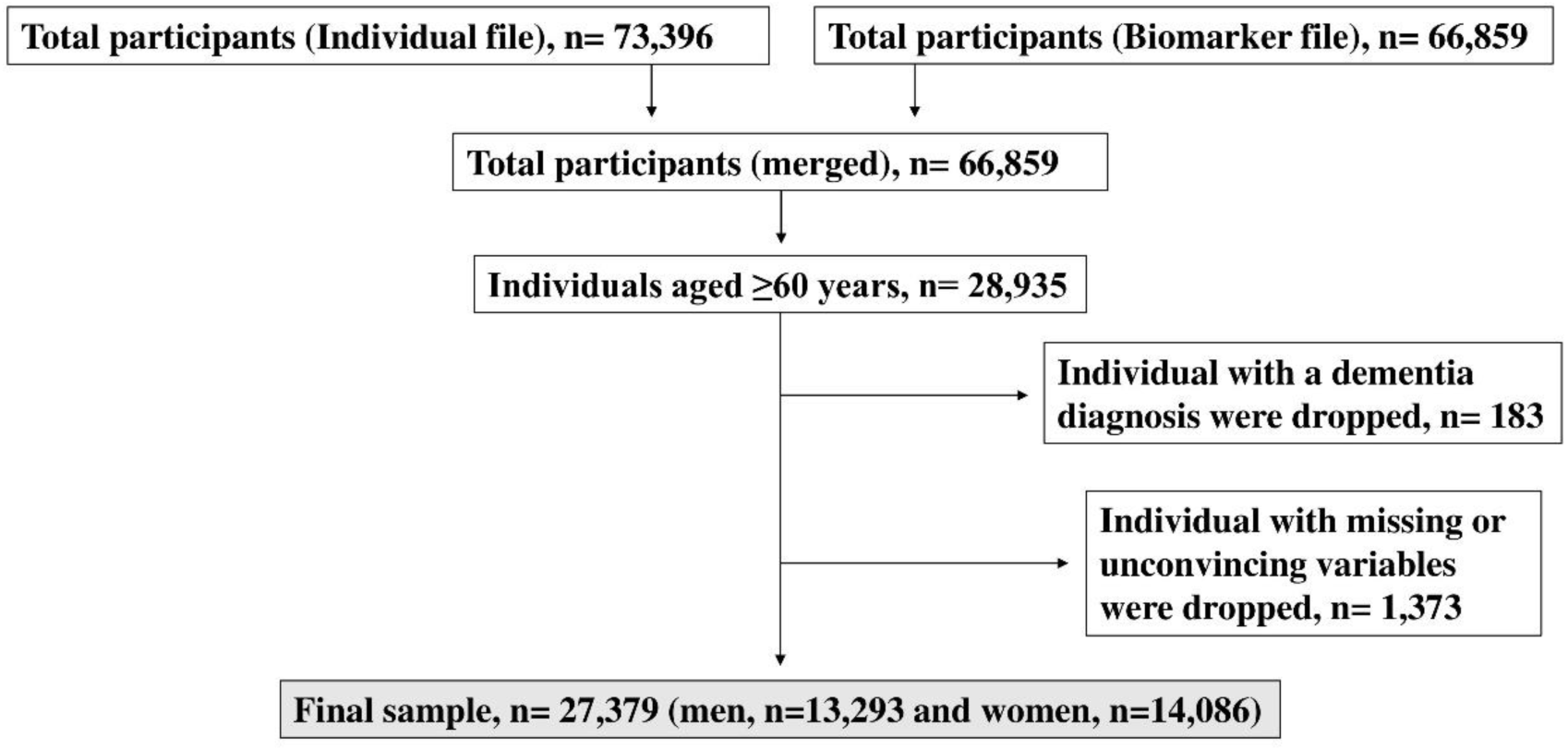
Flow chart of the study sample selection, LASI Wave-1 data

### Outcome variable

The outcome variable for this study was ‘cognitive frailty’ (CF), assessed according to the definition provided by Kelaiditi and colleagues (2013) [16]. In the present study, cognitive frailty was defined as the simultaneous presence of both physical frailty (PF) and cognitive impairment (CI) without dementia.

*Physical frailty (PF)* was evaluated using the modified Community-Oriented Frailty Index (COM-FI) [41], an accumulation of deficits frailty index. We developed a 20-item frailty index based on physical deficits (**Additional File 1**), including i) exhaustion, ii) low body weight (using BMI), iii) low grip strength, iv) low walking speed, v) low physical activity, vi) hypertension, vii) diabetes, viii) lung disease, ix) heart disease, x) stroke, xi) bone problems, xii) neurological problems (but excluding dementia), xiii) polypharmacy, xiv) major injury in the last two years, xv) difficulties in dressing, xvi) difficulties in walking across a room, xvii) bathing difficulties, xviii) eating difficulties, xix) difficulties in getting in and out of bed, and xx) difficulties in using the toilet. Deficits in each domain were scored as 0 (no problem) or 1.0 (problem). Then frailty scores for participants *i*, obtained by summing over the 20 deficits denoted as *j*, were calculated using the following formula, yielding values between 0 and 1.0:

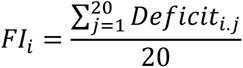

The study categorises scores 0.25 and above as frail and those below 0.25 as non-frail [42]. The assessment of construct validity revealed a Cronbach’s alpha of 0.764 (n=20).

*Cognitive Impairment (CI)* was assessed by evaluating cognitive function, using the composite cognition score based on five cognitive domains. These include i) memory, which involves immediate word recall (0-10) and delayed word recall (0-10); ii) orientation, covering time (0-4) and place (0-4); iii) arithmetic ability, tapping aspects of executive function, evaluated through backward counting from 20 (0-2), a serial 7s’ subtraction task (0-5), and a task involving two computations (0-2); iv) executive functioning and visuospatial functioning skills, measured by paper folding (folding a piece of paper according to instructions) (0-3) and pentagon drawing (drawing intersecting pentagons) (0-1); and finally, v) object naming (0-2). The composite cognitive score ranges from 0 to 43, with a higher value indicating better cognitive function [5, 8]. Additionally, the study also included measures for struggling with instrumental activities of daily living (IADL): i) Preparing a hot meal (cooking and serving); ii) Shopping for groceries; iii) Making telephone calls; iv) Taking medications; v) Doing work around the house or garden; vi) Managing money (paying bills, tracking expenses, etc.); vii) Getting around or finding an address in an unfamiliar place. The composite IADL score ranges from 0 to 7, with a higher value indicating higher disability [43, 44]. To merge both scales, we reversed the score of IADL, and then the total scores ranged from 0 to 50, with higher scores indicating lower cognitive impairment. The lowest 10^th^ percentile was used to indicate cognitive impairment (CI) [40].

*Dementia* assessment was conducted by asking participants about any neurological or psychiatric diagnoses, specifically dementia. If an affirmative response was received, individuals were subsequently excluded from the study based on the cognitive frailty criteria [16].

### Predictor variables

*Sociodemographic factors* included were age (60-69 years, 70-79 years, 80 years and above), gender (man, woman), residence (rural, urban), educational status (less than primary, primary completed, secondary and above), living arrangement (alone, with children, with spouse), and current occupational status (currently working, not working).

The geographical regions were categorised [40] based on all states and UTs in India as follows: North (Jammu and Kashmir, Himachal Pradesh, Punjab, Chandigarh, Uttarakhand, Haryana, Delhi, Rajasthan), Central (Uttar Pradesh, Chhattisgarh, Madhya Pradesh), East (Bihar, West Bengal, Jharkhand, Odisha), Northeast (Sikkim, Arunachal Pradesh, Nagaland, Manipur, Mizoram, Tripura, Meghalaya, Assam), West (Gujarat, Daman and Diu, Dadra and Nagar Haveli, Maharashtra, Goa), and South (Andhra Pradesh, Karnataka, Lakshadweep, Kerala, Tamil Nadu, Pondicherry, Andaman and Nicobar, Telangana).

Religion has been categorised into four major groups: Hindu, Muslim, Christian, and individuals identifying as ‘None of the above,’ reflecting the diverse religious landscape, which also includes Sikh, Buddhist/neo-Buddhist, Jain, Jewish, Parsi/Zoroastrian, and others. This categorisation in the LASI survey is designed to encompass the rich tapestry of religious diversity in the country [40].

Caste is a unique feature in India, where the Government of India recognises four main groups: Scheduled Tribe (ST), Scheduled Caste (SC), Other Backward Class (OBC), and ‘None of the above’ for those who do not belong to any of these categories. This classification is also followed in the LASI survey [40]. ST and SC typically experience more pronounced disadvantages due to historical discrimination, social exclusion, economic and educational disparities. The OBC also face disadvantages compared to the ‘None of the above’ group. However, the extent of these disadvantages varies based on specific contexts, highlighting the importance of recognising and addressing the unique challenges each group faces for inclusive development.

Economic status was assessed based on Monthly Per Capita Consumption Expenditure (MPCE) in Rupees (₹) [40]. Food expenditure was collected based on a reference period of seven days, and non-food expenditure was collected based on reference periods of 30 days and 365 days. Both food and non-food expenditures have been standardised to the 30-day reference period. The MPCE was computed and used as the summary measure of consumption, categorised as (poorest, poorer, middle, richer, richest).

*Physical health and lifestyle characteristics* include self-reported health, tobacco consumption, hospital stays, food insecurity and mobility impairment. Self-Reported Health (SRH) of the respondents was evaluated using the question, “Overall, how is your health in general? Would you say it is very good, good, fair, poor, or very poor?” In our analyses, we utilised a dichotomised version of this variable where ‘poor’ and ‘very poor’ were coded as “poor,” and ‘very good,’ ‘good,’ and ‘fair’ were coded as “good” for poor SRH [45].

Tobacco consumption was assessed with the question, “Have you ever smoked tobacco (cigarette, bidi, cigar, hookah, cheroot) or used smokeless tobacco (such as chewing tobacco, gutka, pan masala, etc.)?”, the responses were dichotomous, recorded as yes or no [46].

Hospital stays were identified based on the question, “Over the last 12 months, how many times were you admitted as a patient to a hospital/long-term care facility for at least one night?” If the response was one or more times, it was categorised as yes; otherwise, it was categorised as no [47].

Food insecurity was measured using four questions: (a) In the last twelve months, did you ever reduce the size of your meals or skip meals because there was not enough food in your household? (b) In the last twelve months, were you hungry but didn’t eat because there was not enough food in your household? (c) In the past twelve months, did you ever not eat for a whole day because there was not enough food in your household? (d) Do you think that you have lost weight in the last twelve months because there was not enough food in your household? The present study considered respondents who answered ‘yes’ to at least one of these questions to be ‘food insecure’ [48].

Mobility impairment was assessed based on various activities, such as (a) walking 100 yards, (b) sitting for 2 hours or more, (c) getting up from a chair after sitting for a long period, (d) climbing one flight of stairs without resting, (e) stooping, kneeling or crouching, (f) reaching or extending arms above shoulder level, (g) pulling or pushing large objects, lifting or carrying weights over 5 kilos like a heavy bag of groceries, and (h) picking up a coin from a table. The responses for these questions were recorded as ‘no’ or ‘yes.’ The scale was summed up and recoded into a dichotomous measure as ‘No’ for no mobility difficulty and ‘Yes’ otherwise [49].

*Psychosocial health characteristics* include sleep problems, major depression, social participation, life satisfaction and everyday discrimination. Sleep problems were assessed using four questions adapted from the Jenkins Sleep Scale (JSS-4): (a) How often do you have trouble falling asleep? (b) How often do you have trouble waking up during the night? (c) How often do you have trouble waking up too early and not being able to fall asleep again? (d) How often did you feel unrested during the day, no matter how many hours of sleep you had? Response options included ‘never,’ ‘rarely’ (1–2 nights per week), ‘occasionally’ (3–4 nights per week), and ‘frequently’ (5 or more nights per week) (item d was reverse coded). Sleep problems were coded as “Yes” if any of the four symptoms occurred frequently [50]. The scale demonstrated excellent reliability (Cronbach α= 0.873, n=4).

Major depression was assessed using the Composite International Diagnostic Interview short-form (CIDI-SF) scale, consisting of ten questions. Responses, except for items two and three, were binary (“No” coded as 0 and “Yes” coded as 1). Individuals who felt sad, blue, or depressed ‘all day long’ or ‘most of the day’ were coded as “Yes”; similarly, those who felt sad, blue, or depressed ‘every day’ or ‘almost every day’ were coded as “Yes”. The CIDI-SF depression scale, ranging from 0 to 10, categorised respondents with a score of 5 and more as “Depressed” and those with a score of 4 and less as “Not depressed” [51].

Social participation was assessed based on engagement in social activities, including (a) eating out, (b) going to parks/beaches, (c) visiting relatives/friends, (d) attending cultural performances/shows/cinema, (e) participating in religious functions/events, and (f) attending community/political/organisation group meetings. Respondents reporting participation in any of these activities at least once a month were considered to have social participation (1 = at least once a month, 0 = rarely or never) [52] (Cronbach’s alpha: 0.677, n=11).

Life satisfaction was evaluated using statements regarding overall life contentment. Responses were categorised as ‘strongly disagree,’ ‘somewhat disagree,’ ‘slightly disagree,’ ‘neither agree nor disagree,’ ‘slightly agree,’ ‘somewhat agree,’ and ‘strongly agree.’ Scores ranged from 5 to 35, with higher scores indicating greater life satisfaction. The scale was further categorised into tertiles: “low satisfaction” (5–20), “medium satisfaction” (21–25), and “high satisfaction” (26–35) [53]. This scale demonstrated excellent reliability (Cronbach α= 0.899, n=5).

Everyday discrimination experiences were assessed with the six-item Everyday Discrimination Scale (Cronbach’s alpha: 0.862, n=6). Responses ranged from 1 = ‘Almost every day’ to 6 = ‘never’ and were dichotomised into ‘never’ = “no discrimination” and ‘ever’ (collapsing those reporting ‘less than once a year’ or greater into one category) = “high discrimination” [50].

### Statistical analysis

Descriptive statistics were utilised to present a comprehensive overview of the background characteristics within the study population, categorised by CF status. To determine differences in category membership between individuals with and without CF, chi-square test (χ2 test) was conducted for all categorical variables. At the initial stage, variables where the χ2 tests had p-values <0.01 were included in the binary logistic regression analysis. To explore the association between each predictor variables and the outcome (CF) variable individually, the study estimated unadjusted odds ratios through binary logistic regression. Subsequently, in order to identify a significant proportion of the variance in the outcome and predictor variables, stepwise binary logistic regression (backward elimination) was executed. This approach, following the method outlined by Hosmer and Lemeshow [54], provided adjusted odds ratios (AOR) to better understand the associations.

When considering the predictor variable *X* with a binary (categorical) outcome variable *Y*, the logistic regression equation is expressed as:

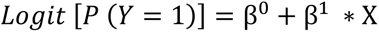

In this equation, the logit function, denoted as *Logit[P(Y=1)]*, transforms the probability of the event *Y=1* into a scale ranging from negative to positive infinity. *β^0^* is the intercept term which estimates the log odds of the event *Y=1* when all predictor variables (*X*) are equal to zero, and *β^1^* is the coefficient associated with the predictor variable *X*, estimates the change in the log odds of the event *Y=1* for a one-unit change in the predictor variable X. Through the maximum likelihood estimation, *β^0^* and *β*^1^ are derived from the data, providing insights into the log odds and the impact of the predictor variable *X* on the log odds. This helps quantify the differential log odds of the outcome variable associated with the set of predictors (*X*) compared to the reference group. Variables demonstrating a significant χ2 (p < 0.01) were then classified into three distinct groups: (1) sociodemographic, (2) physical health and lifestyle characteristics, and (3) psychosocial characteristics associated with cognitive frailty. Following this, a series of logistic regression analyses were conducted, with the three categorised groups as criteria. Variables demonstrating a significant contribution (p < 0.01) in each model were selected for inclusion in the final logistic model. The study reported adjusted odds ratios (AOR) and their corresponding 95% confidence intervals, the 2 log-likelihood statistic, Pearson’s χ^2^, and Nagelkerke’s R^2^ as indicators of model fit. All statistical analyses were performed using IBM Statistical Package for Social Sciences (IBM SPSS version 22.0, SPSS Inc., Chicago, IL, USA) software.

## Results

### Prevalence of cognitive frailty in India by region

In **Figure 2**, the Q-GIS (Quantum Geographic Information System) offers a depiction of regional variations in the prevalence of CF in India based on a sample of 27,379 (N) participants. High prevalence is evident in the Western region, where all states exhibit a prevalence higher than the overall national prevalence of 4.5% (n=1243) (**Table 1**). Specifically, Goa (7.8%), Dadra and Nagar Haveli (7.8%), Maharashtra (6.6%), Gujarat (4.7%), and Daman and Diu (4.5%) show elevated rates. Southern states also demonstrate considerable prevalence, including Telangana (7.1%), Lakshadweep (6.3%), Andaman and Nicobar (6.2%), Andhra Pradesh (5.7%), Karnataka (5.6%), Kerala (5.1%), and Tamil Nadu (4.6%). In the Eastern part, West Bengal reports a notably high prevalence of 7.1%, while in the Northern region, Jammu and Kashmir (7.0%) and Tripura in the Northeast (5.2%) show elevated rates. It is noteworthy that almost all Northeast states exhibit lower prevalence than the national average, with Nagaland reporting the lowest prevalence at 0.7%

**Figure 2.**
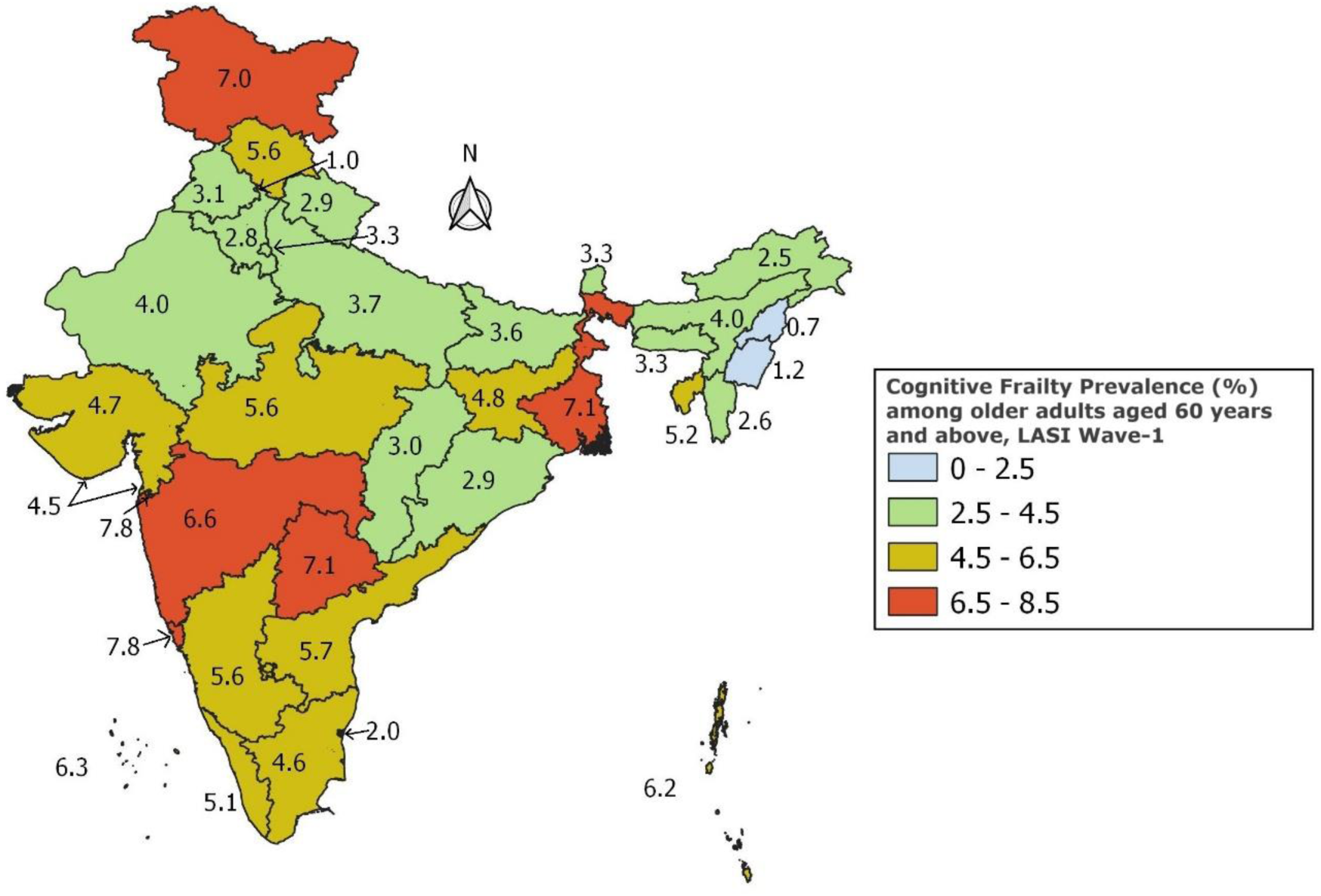
Prevalence of Cognitive Frailty among older adults aged 60 years and above in India by its states, LASI Wave 1 data

**Table 1.** Sociodemographic characteristics of the study population in India, LASI Wave-1 (N=27,379)

|  |  |  |  |  |  |
| --- | --- | --- | --- | --- | --- |
| **Sociodemographic characteristics** |  | **Cognitive Frailty, n (%)** |  | **$\chi^2$ test value** | **p-value** |
|  |  | **Yes**<br>**(n=1243, 4.5%)** | **No**<br>**(n=26136, 95.5%)** |  |  |
| Age | 60-69 | 343 (2.0) | 16675 (98.0) | 1059.09 | <0.01 |
|  | 70-79 | 500 (6.4) | 7343 (93.6) |  |  |
|  | 80+ | 400 (15.9) | 2118 (84.1) |  |  |
| Gender | Man | 298 (2.2) | 12995 (97.8) | 314.89 | <0.01 |
|  | Woman | 945 (6.7) | 13141 (93.3) |  |  |
| Region | North | 190 (3.8) | 4814 (96.2) | 64.73 | <0.01 |
|  | Central | 154 (4.2) | 3553 (95.8) |  |  |
|  | East | 234 (4.6) | 4865 (95.4) |  |  |
|  | Northeast | 101 (2.8) | 3473 (97.2) |  |  |
|  | West | 227 (6.2) | 3408 (93.8) |  |  |
|  | South | 337 (5.3) | 6023 (94.7) |  |  |
| Residence | Urban | 268 (3.0) | 8775 (97.0) | 77.42 | <0.01 |
|  | Rural | 975 (5.3) | 17361 (94.7) |  |  |
| Religion | Hindu | 914 (4.6) | 19085 (95.4) | 25.57 | <0.01 |
|  | Muslim | 186 (5.9) | 2993 (94.1) |  |  |
|  | Chirstian | 90 (3.3) | 2675 (96.7) |  |  |
|  | None of the above | 53 (3.7) | 1383 (96.3) |  |  |
| Caste | Scheduled tribe | 239 (5.2) | 4387 (94.8) | 24.09 | <0.01 |
|  | Scheduled caste | 243 (5.5) | 4198 (94.5) |  |  |
|  | Other backward caste | 466 (4.4) | 10015 (95.6) |  |  |
|  | None of the above | 295 (3.8) | 7536 (96.2) |  |  |
| Educational status | <Primary | 1185 (6.6) | 16818 (93.4) | 509.26 | <0.01 |
|  | Primary completed | 51 (1.0) | 5278 (99.0) |  |  |
|  | Secondary and above | 7 (0.2) | 4070 (99.8) |  |  |
| Living arrangement | Alone | 93 (6.6) | 1320 (93.4) | 602.70 | <0.01 |
|  | With children | 758 (8.9) | 7737 (91.1) |  |  |
|  | With spouse | 292 (2.2) | 17079 (97.8) |  |  |
| Current occupational status | Working | 80 (0.9) | 8378 (99.1) | 364.80 | <0.01 |
|  | Not working | 1163 (6.1) | 17758 (93.9) |  |  |
| Economic status | Poorest | 319 (5.7) | 5288 (94.3) | 34.41 | <0.01 |
|  | Poorer | 264 (4.7) | 5399 (95.3) |  |  |
|  | Middle | 266 (4.7) | 5383 (95.3) |  |  |
|  | Richer | 220 (4.1) | 5190 (95.9) |  |  |
|  | Richest | 174 (3.4) | 4876 (96.6) |  |  |

### Sociodemographic characteristics

The sociodemographic characteristics of the study population in India (N=27,379) were investigated concerning CF status (**Table 1**). The geographical regional disparities showed statistically significant differences, with the highest prevalence observed in the Western region (6.2%, n=227), p < 0.01) followed by the Southern region (5.3%, n=337), p < 0.01) and Eastern region (4.6%, n=234), p < 0.01). The prevalence of CF exhibited an increasing trend with age, particularly notable in the 70-79 age group (prevalence = 6.4%, (n=500), p < 0.01) and the 80+ age group (15.9%, (n=400), p < 0.01). Gender disparities were apparent, with women demonstrating a higher prevalence (6.7%, (n=945), p < 0.01). Urban-rural residence emerged as a noteworthy factor, with rural areas displaying higher rates of CF (5.3%, n=975), p < 0.01). The prevalence of CF is higher among individuals belonging to the Muslim religion (5.9%, n=186, p <0.01), and among the SC (5.5%, n=243, p <0.01) as well as the ST (5.2%, n=239, p <0.01) within the caste category, compared to their respective counterparts. Educational status revealed a robust association, with lower education linked to an increased prevalence of CF (6.6%, n=1185, p < 0.01). Living with children (8.9%, n=758), or alone (6.6%) were higher CF than living with spouse (2.2%) (p < 0.01). Currently not working (6.1%, n=1163), p < 0.01), and poorest economic status (5.7%, n=319), p < 0.01) were all significantly associated with CF status (p<0.01), highlighting the intricate interplay of sociodemographic factors with CF in the older Indian population.

### Physical health, lifestyle and psychosocial characteristics

**Table 2** provided a comprehensive examination of the physical health, lifestyle, and psychosocial wellbeing characteristics of the study population in India (N=27,379) concerning CF status. A robust association was observed between SRH status and CF (p < 0.01), with 10.8% (n=633) of individuals reporting poor health being in the CF category and only 2.8% of those with good health showing CF. There was no observed association between current smoking status and CF (p = 0.154). Hospital stays in the last 2 years (p < 0.01), food insecurity (p < 0.01), and mobility impairment (p < 0.01) were all significantly associated with CF. In psychosocial health, sleep problems, social participation, low life satisfaction, and everyday discrimination (all p < 0.01) showed strong associations with CF status, with CF being more likely in those with each negative issue. Major depression was also significantly associated with CF (p < 0.01), with 11.0% (n=203) of individuals with depression being in the CF category and only 4.1% of those without depression.

**Table 2.** Physical health, and lifestyle and psychosocial characteristics of the study population in India, LASI Wave-1 (N=27,379)

| Physical health and lifestyle characteristics | | Cognitive Frailty | | $\chi^2$ test value | p-value |
| --- | --- | --- | --- | --- | --- |
|  |  | Yes<br>(n=1243, 4.5%) | No<br>(n=26136, 95.5%) |  |  |
| Self-reported health status | Good | 610 (2.8) | 20910 (97.2) | 674.86 | <0.01 |
|  | Poor | 633 (10.8) | 5226 (89.2) |  |  |
| Tobacco use (ever) | Yes | 465 (4.3) | 10316 (95.7) | 2.11 | 0.154 |
|  | No | 778 (4.7) | 15820 (95.3) |  |  |
| Hospital stays in last 2 years | Yes | 158 (7.7) | 1885 (92.3) | 51.96 | <0.01 |
|  | No | 1085 (4.3) | 24251 (95.7) |  |  |
| Food insecurity | Yes | 201 (8.5) | 2163 (91.5) | 93.74 | <0.01 |
|  | No | 1042 (4.2) | 23973 (95.8) |  |  |
| Mobility impairment | Yes | 1213 (6.1) | 18651 (93.9) | 409.80 | <0.01 |
|  | No | 30 (0.4) | 7485 (99.6) |  |  |
| Psychosocial health |  |  |  |  |  |
| Sleep problem | Yes | 292 (9.8) | 2678 (90.2) | 215.25 | <0.01 |
|  | No | 351 (3.9) | 23458 (96.1) |  |  |
| Major depression | Yes | 203 (11.0) | 1647 (89.0) | 189.46 | <0.01 |
|  | No | 1040 (4.1) | 24489 (95.9) |  |  |
| Low social participation | Yes | 545 (10.0) | 4913 (90.0) | 466.41 | <0.01 |
|  | No | 698 (3.2) | 21223 (96.8) |  |  |
| Life satisfaction | Low | 547 (6.7) | 7606 (93.3) | 131.47 | <0.01 |
|  | Medium | 268 (4.1) | 6257 (95.9) |  |  |
|  | High | 428 (3.4) | 12273 (96.6) |  |  |
| Everyday discrimination | Yes | 284 (6.6) | 3999 (93.4) | 51.22 | <0.01 |
|  | No | 959 (4.2) | 22137 (95.8) |  |  |

### Association of sociodemographic characteristics, physical health, lifestyle and psychosocial health characteristics with CF status

**Table 3** showed the unadjusted and adjusted odds ratios explaining the associated factors influencing CF in the study population. The unadjusted models underscore significant associations between advanced age and CF status, with individuals aged 70-79 years exhibited OR of 3.3 (95% CI 2.9-3.8, p < 0.01), and those aged 80 and above displayed a notably higher OR of 9.2 (95% CI 7.9-10.7, p < 0.01), with age 60-69 being the referent. Women (OR 3.1, 95% CI 2.7-3.6, p < 0.01), residence in rural areas (OR 1.8, 95% CI 1.6-2.1, p < 0.01), and specific geographical regions, such as the West (OR 1.7, 95% CI 1.4-2.1, p < 0.01) and South (OR 1.4, 95% CI 1.2-1.7, p < 0.01), were also significantly associated with increased odds of CF. Religious affiliations such as Muslims, specific caste categories like the more disadvantaged castes: SC, ST and OBC, lower educational attainment, living arrangements of living alone or with children, current occupational status of not working, and the poorest economic status all exhibited significant associations (p < 0.01) with increased odds of CF. Additionally, poor SRH status, hospital stays, food insecurity, mobility difficulties, and poor psychosocial health variables also demonstrated significant associations with CF (p < 0.01).

**Table 3.**
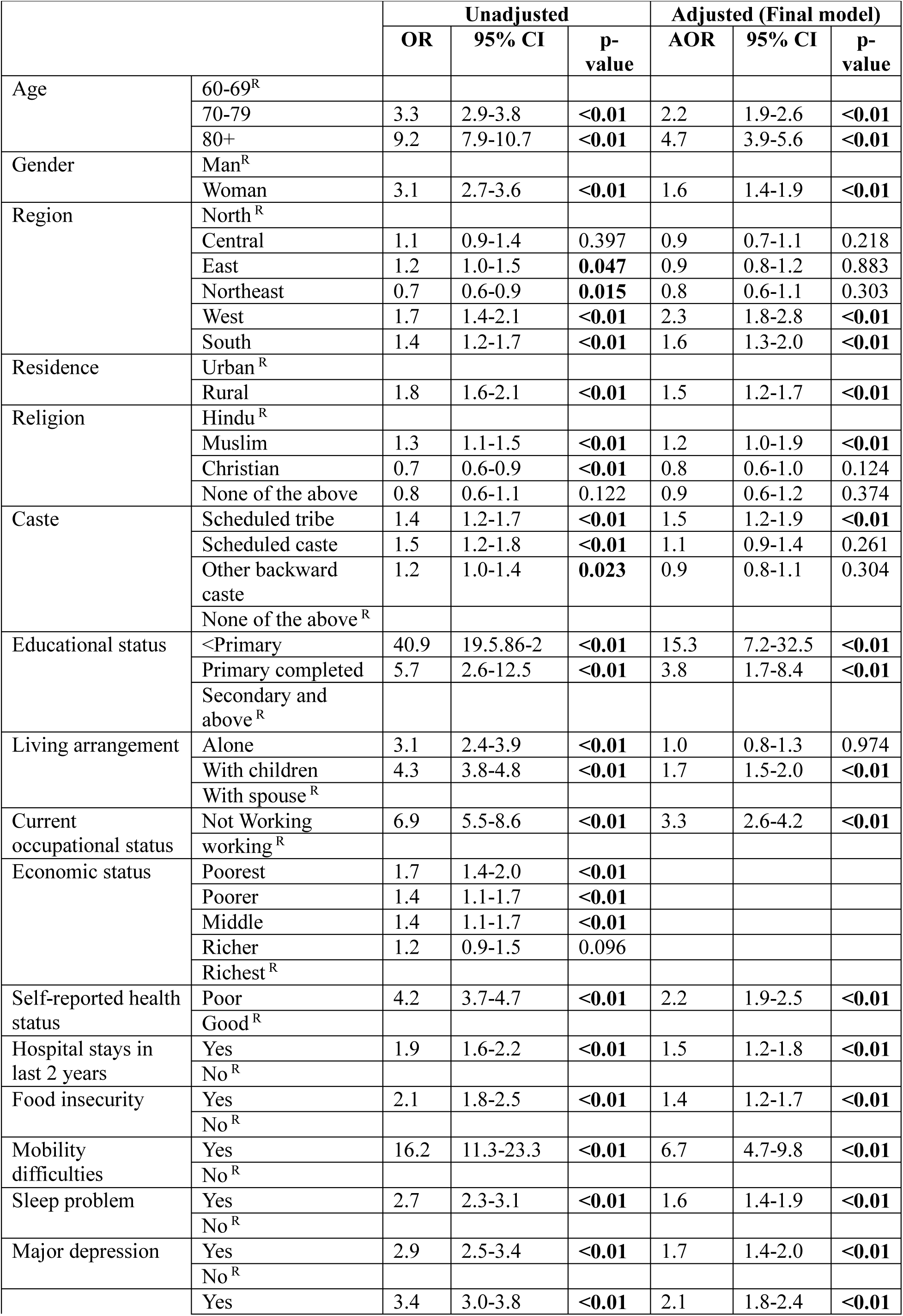
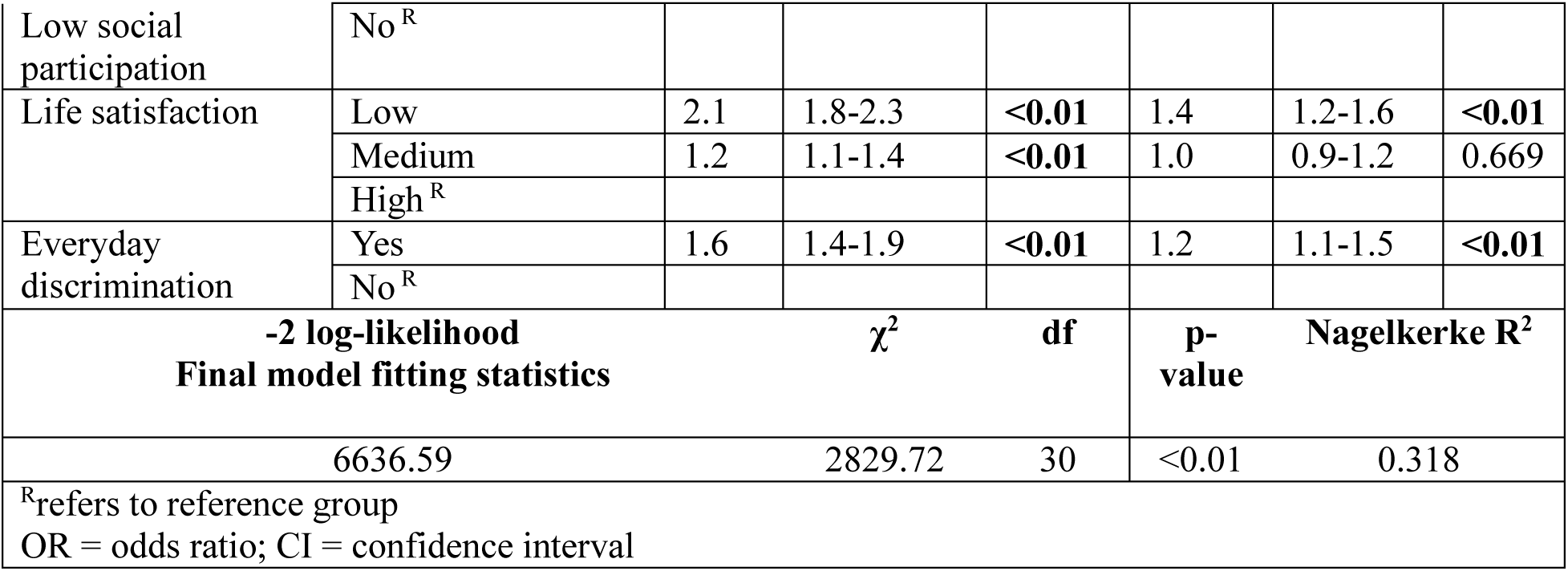
Result of stepwise (backward elimination) binary logistic regression analysis (N=27,379)

In the final model (adjusted for all sociodemographic characteristics except economic status, physical and lifestyle characteristics and psychosocial health), several significant predictors of CF were identified. Age displayed a graded effect, with the 70-79 age group showing an adjusted odds ratio (AOR) of 2.2 (95% CI: 1.99–2.6, p < 0.01) and the 80+ age group exhibiting the highest AOR of 4.7 (95% CI: 3.9–5.6, p < 0.01) compared to the reference group (60-69). Women was associated with a higher likelihood of CF (AOR = 1.6, 95% CI: 1.4–1.9, p < 0.01). Regional variations were observed, with individuals from the West (AOR = 2.3, 95% CI: 1.8–2.8, p < 0.01) and South (AOR = 1.6, 95% CI: 1.3–2.0, p < 0.01) demonstrating higher odds. Other significant predictors included rural residence (AOR = 1.5, 95% CI: 1.2–1.7, p < 0.01), educational status below primary level (AOR = 15.3, 95% CI: 7.2–32.5, p < 0.01), currently not working (AOR = 3.3, 95% CI: 2.6–4.25, p < 0.01), poor self-reported health (AOR = 2.2, 95% CI: 1.9–2.5, p < 0.01), and mobility impairment (AOR = 6.7, 95% CI: 4.7–9.8, p < 0.01). Hospital stays in last 2 years (AOR = 1.5, 95% CI: 1.2–1.8, p < 0.01), and food insecurity (AOR = 1.4, 95% CI: 1.2–1.7, p < 0.01) also exhibited high odds for CF status. Psychosocial factors such as sleep problems (AOR = 1.6, 95% CI: 1.4–1.9, p < 0.01), major depression (AOR = 1.7, 95% CI: 1.4–2.0, p < 0.01), low social participation (AOR = 2.1, 95% CI: 1.8–2.4, p < 0.01), and low life satisfaction (AOR = 1.4, 95% CI: 1.2–1.6, p < 0.01), also demonstrated significant associations with cognitive frailty. To achieve an optimal fit, the final model, employing stepwise (backward elimination) binary logit model approach, eliminated economic status from the set of predictor variable (which was not significant after adjustment). The final model fitting statistics, including the −2 log-likelihood, χ2, degrees of freedom (df), and Nagelkerke R^2^, collectively indicate the robustness of the model in explaining the variance in CF, offering valuable insights for targeted interventions and holistic healthcare strategies.

## Discussion

The first aim of this study was to examine the national prevalence of CF in India. While increased longevity in India is to be celebrated, the rapid increase in the older population in India with a projected population of 347 million over 60s by 2050 [1], brings an urgent need for understanding the current and potential health status and care needs of the older population. This understanding is crucial to develop policy and public health strategies with the purpose of reducing prevalence of serious late-life impairments. Developing understanding and risks for CF is an important part of this, as is development of potential interventions for this reversible [21, 36, 37], but high-risk condition [22–24, 31, 35]. In a descriptive review, Sugimoto and colleagues (2018) [55] demonstrated a prevalence of 1.0-1.8% of CF in community-based samples, with higher levels in clinical samples (e.g. in participants recruited from specific disease groups or hospital frailty units). They also noted higher levels when neurodegenerative conditions such as dementia were not excluded, and that levels increased with the age of the sample. In a meta-analysis, Qiu and colleagues (2021) [56] carefully excluded studies that did not exclude participants with dementia, and showed that pooled prevalence of CF was higher, at 9%, although this study did include studies with higher age ranges (e.g. one study only considered participants aged over 90 years). However, no prevalence review or meta-analysis could be found that included any studies from India, with only one previous study [23] estimating prevalence in one region of India amongst a rural population (West Bengal) as 21.8% amongst those aged over 60 years. This study found a national prevalence of CF of 4.5% which is higher than many previous studies based on the general over 60s population [55].

In response to our second aim, the present study also demonstrated significant variance by geographical region of India with the highest prevalence of CF in Western and Southern states. On a local level within these regions, some areas were noted as having prevalence rates of over 6.0%, surpassing the national prevalence of 4.5%. It is already well-established that South India has become a major driver of the ageing population in India, compared to other regions, with an increasing trend in the burden of geriatric health problems [1, 57–60]. A recent population-based study from the state of Telangana reported that every fifth older adult had at least one disability, and every third individual had at least one non-communicable disease (NCD) [61]. In our study, this state showed the highest prevalence of CF among Southern India, at 7.1%. It is also reported that, compared to older adults in Southern India, those in Northern, Eastern, North-Eastern, and Western India had lower levels of other psychosocial health issues like loneliness [62]. On the other hand, Patel and colleagues (2023) [63] highlighted a high burden of multimorbidity in both Western and Southern India, emphasising the need for increased attention to these states in the country. In our study, the Western state of Goa showed a high prevalence of CF (7.8%) in this region. Goa is second, in the ‘old age dependency’ ratio in India, to Kerala, a Southern state [40]. Additionally, Patel and Prince’s qualitative study (2001) [64] on mental health needs among the aged in Goa revealed that, despite being frequently observed in the older population in this state, several mental health deficits were still not acknowledged as health needs. The regional variation in CF prevalence in the present study suggests a need for a significant focus on these regions, considering the upcoming demographic transition. Given that our findings indicate higher rates of CF among those living in rural communities, among those with lower socioeconomic resources, lower education and of specific religious affiliation, it is suggested that these regions may also differ from those with lower prevalence of CF in these characteristics. Therefore, it is suggested that the relatively high prevalence of CF amongst the older population in India is related to the high frequency of these risk factors.

The third aim of this study was to consider the association of CF with potential risk factors, using a holistic approach that includes a range of intrinsic and extrinsic variables. The study first examined socio-demographic variables such as age and gender, confirming expectations of an increase in prevalence with age (e.g. to 15.9% of the over 80s) in this age related condition, although this was still lower than the prevalence of 50.3% found in Hao and colleagues’ (2018) [65] study of people aged over 90 years in the Sichuan region of China, and lower than the prevalence of 43.9% found amongst over 80s in the smaller study by Navarro-Pardo and colleagues (2020) [32] in Spain. The present study also confirmed previously noted disparities between men and women, with women showing a 6.7% prevalence of CF compared to 2.2% in men and an odds ratio indicating they were 3.1 times more likely to be cognitively frail than men, although this reduced to 1.6 times in the adjusted model. Higher frequency of CF in older women has been related to longer life expectancy in women, to reduced opportunities for education and occupational or social roles (reducing lifetime intellectual stimulation and so cognitive reserve) [66–68], as well as to differential underlying biological changes related to hormonal changes [69].

The effect of having less than primary education (6.6% CF prevalence) was striking as compared with having secondary education (0.2% CF) and showing an adjusted odds ratio of 15.3 times the risk of CF than in those with secondary education or above. As the strongest effect in the study, this suggests that policies that increase the proportion of people with secondary education or higher in India could gradually have a very significant impact on the health of people in older age. From the data in the LASI dataset, it can be seen that more than 60.0% of over 60s have less than primary education [70, 71]. World bank data (2011) [72] indicates that only 40% of Indian adolescents attend secondary school, but 95% of younger children are now attending primary school. While there are many economic reasons for increasing the educational level of a population, present study shows the long-term impacts on the health, independence and potential care needs of India’s ageing population.

While economic status based on income did not show any association with CF in the adjusted model, food insecurity showed consistent increased odds of CF including in the adjusted model. Mechanisms by which poverty and inequalities impact on likelihood of CF in older age include impact on potential nutritional deficiencies. Poor or restricted diet and particularly low protein intake in older adults can lead to weight loss and reductions in muscle mass, both indicators of PF. Links between nutrition and cognition have been well researched, with several previous studies providing evidence of a link with both PF and CI [73, 74], and lower consumption of some nutrient groups including whole grains, vegetables, fruit, meat and nuts. In addition, low vitamin D and omega 3 polyunsaturated fatty acids has been associated with coexisting PF and CI [75]. A previous study using the LASI data [76] found that food security was strongly associated with cognitive function in terms of word recall and the arithmetic tests described in this study, with those older adults who did have food security being 0.71 and 0.45 times less likely (respectively) to show impairment in these cognitive tests than those who reported food insecurity. Other studies have shown a strong link between food insecurity and cognitive decline in older age; for example, a longitudinal study in a Puerto Rican population in the United States showed, in a fully adjusted model, that food insecurity was associated with a faster decline in executive function (but not memory) [77]. The distinction between executive function and memory is important as CF is commonly associated with the former more than the latter [78].

Another potential impact of inequalities examined was the impact of experiences of everyday discrimination. While this may also be associated with religious affiliation, caste or poverty which separately impacted odds for CF, this variable indicated one mechanism whereby issues like religious affiliation and caste may have an impact on wellbeing. One of the key biological mechanisms suggested for this co-occurrence of PF and CI is the underlying changes in the immune system (immunosenescence), in inflammation (inflammageing) and in the gut microbiome, all of which can be affected by long term stress, including stresses such as repeated social defeat stress [79].

Psychological wellbeing was also considered: depression, sleep disorder and life satisfaction all showed significant effects on adjusted odds of CF. While again these issues could all be bidirectional, with CF resulting in depression or lower life satisfaction, previous longitudinal studies have specifically demonstrated the role of depression as a mediator in the relationship between baseline frailty and later cognitive impairment [80], highlighting it as a potential important target for intervention.

Living with one’s children and not working were also associated with a higher adjusted odds ratio of CF, although both may also be associated with other aspects of ageing such as retirement. Living with one’s children or having to stop working may not necessarily be a risk factor, but rather a consequence of existing higher CF. However, an indicator of social support and also socially and intellectually stimulating activity can be seen in the measure of social participation, which included a range of activities. In the adjusted model, lower social participation was associated with increased odds of cognitive frailty. Mechanisms of the effects of social participation include strengthening of cognitive reserve, particularly important in a population with low education or lower-level occupations, but also impacts on loneliness and availability of support in stressful situations. People who are more socially active outside the home are also more likely to be more physically active, reducing or ameliorating the impact of physical frailty on cognition [22, 81], and potentially impacting nutrition by being more likely to eat with other people.

Self-reported poor health status, having had a hospital stay and experiencing mobility difficulties were all associated with odds for CF although one may argue that they may also be consequences of CF. It must be borne in mind that this is a cross-sectional study, given that the LASI has not yet published the second wave of data collection. Future analyses could examine this question of direction of association by examining risk of occurrence of CF at Wave 2 in those who were not living with CF at Wave 1.

Although a large proportion of this population reported that they had used tobacco (around 30%), this included people who had smoked it and used it in other ways such as chewing tobacco. The question was also whether they had ever used it, as opposed to current or recent use or amount of use, so while tobacco smoking is well-evidenced as a health risk and as a risk for dementia, it did not show in this study as affecting risk of CF.

### Strengths and Limitations

The key strength of this study is that it is the first analysis of the prevalence and risk factors of cognitive frailty in India, providing a comprehensive overview in a very varied population, representing the highest national population in the world. The study presents a range of prevalence data for different population and regional variabilities, all of which can give a useful basis for more in-depth studies within specific regions or populations groups in the future.

The most salient limitation of this study is that it is cross-sectional rather than longitudinal. Future waves of the LASI will enable predictive models to be refined and directionality to be investigated in terms of the development and trajectory of CF. The data available are specific to India which means that comparisons with other datasets are not necessarily appropriate – for example, the ethnic and religious mix, regional, cultural, educational and economic differences, and caste identity are unique to India. While other studies have compared CF prevalence in regions of a country with complex regional and ethnic differences within a large population, and confirmed, as here, differences in prevalence of CF, only some of the factors may be comparable, for example, differential access to education or other inequalities [82]. This may be a limitation in terms of comparability but is a strength in terms of beginning to understand some of the predictors and potential intervention targets of CF in the older Indian population, which, as indicated, is essential as the older population grows so substantially.

The range of measures available for assessing CF provided some limitations. It was relatively easy to construct a frailty index based on previously validated accumulation of deficits indices, but the range of cognitive measures did not match well to those used in previous studies. Many studies use a cut-off on the Mini-Mental State Examination (MMSE) to determine cognitive impairment for CF, but that is increasingly seen as inappropriate given evidence for the importance of executive function in CF and the lack of clear indicators of executive function in the MMSE. The Montreal Cognitive Assessment (MoCA) or other global cognitive measures that include executive function is more useful in this context and is now commonly used. However, few large cohort datasets include it, and most researchers use a mix of individual indices. Our mix was based on that used by Muhammad and colleagues (2023) [8] and measures in the LASI were based on MMSE components, among other original measures and the use of visuospatial function indicators for executive function could be updated in future studies. However, as the measures in LASI included an “arithmetic” category that seem to tap into aspects of executive functioning such as updating, the cognitive composite score was seen to be useful.

### Implications for policy and practice

The variables that had the most salient impacts on odds of CF include several factors that are modifiable. Educational background had the biggest impact of all the variables examined and may also be related to some of the other regional, economic, ethnic and gender disparities [83]. While increasing the proportion of the population that completes secondary education seems urgent, the mechanism by which education impacts later life cognitive health includes the impact on cognitive reserve, a factor that can delay the impact of any neurodegeneration. Cognitive reserve can be built in many ways throughout life in addition to early life education, and so making opportunities for occupational learning and intellectually stimulating leisure activities is also important.

Social participation was another salient factor that increased the odds of being in the CF group. Research that examines the causes and interventions for social isolation and loneliness has largely been done in Western countries and although there will be similarities, research specifically in the different cultures represented in India’s regions is needed.

Finally, food insecurity is a policy issue. The World Food Programme states that India is home to one quarter of the world’s undernourished people, and amongst older people, inadequate food intake and vitamin deficiencies are associated with increased likelihood and earlier significant cognitive impairment [84]. Focus on the nutrition of India’s older population is an important focus [85].

## Conclusions

This is the first study to describe the prevalence of cognitive frailty across India, showing higher prevalence than may be expected based on global figures for the general over 60s community population, with clear regional disparities. The study examined a range of risk factors related to both individual factors such as depression, life satisfaction and social participation, as well as a range of sociocultural and socioeconomic determinants such as level of education, experiences of discrimination, income and food insecurity. The study highlighted the impact of several of these in an adjusted model and considered potential implications for policy. Future availability of longitudinal data and specific in-depth investigations will be important to build on this work.

## Supporting information

. We developed a 20-item frailty index based on physical deficits (Additional File 1),

## Data Availability

The datasets analysed in this study are accessible through the Institute for Population Sciences (IIPS) Mumbai repository. IIPS Mumbai served as the designated entity for data collection on behalf of the Ministry of Health and Family Welfare, Government of India. Researchers and policymakers can obtain the data and materials of the LASI Wave-1 by formally requesting access from the IIPS. The necessary information and request form are available through the provided links: https://www.iipsindia.ac.in/content/LASI-data

## List of abbreviations

CEB: Census Enumeration Block
CF: Cognitive frailty
CI: Cognitive impairment
CIDI-SF: Composite International Diagnostic Interview short-form
COM-FI: Community-Oriented Frailty Index
IADL: Instrumental Activities of Daily Living
LASI: Longitudinal Ageing Study in India
MMSE: Mini-Mental State Examination
MoCA: Montreal Cognitive Assessment
MPCE: Monthly Per Capita Consumption Expenditure
OBC: Other Backward Class
PF: Physical Frailty
PSUs: Primary Sampling Units
SC: Scheduled Caste
SRH: Self-Reported Health
ST: Scheduled Tribe
UNFPA: United Nations Population
Fund UTs: Union Territories

## Ethics approval and consent to participate

The LASI Wave 1 survey had received ethical approval from the Indian Council of Medical Research (ICMR), India. All research methods were conducted in strict adherence to the pertinent guidelines and regulations. Informed consent was obtained from all subjects. Additionally, necessary ethical clearance for using secondary data was obtained from the Lancaster University Faculty of Health and Medicine Ethics Committee (FHMREC), UK with the ethics reference FHM-2023-3659-DataOnly-1.

## Consent for publication

Not applicable for this study.

## Authors’ contributions

SD and CH conceptualised the idea and jointly drafted the manuscript; SD and EP conducted the statistical analysis. SD and CH critically reviewed and made contributions to the initial manuscript. The final manuscript has been read and approved by all authors.

## Acknowledgements

Author acknowledges the role of International Institute for Population Sciences (IIPS) website for providing the secondary data with a request for use in research purposes only. The authors extend their sincere appreciation to Dr Sutapa B. Neogi, Director, and Dr Preetha G. S., Research Dean of the International Institute of Health Management Research (IIHMR), New Delhi, India, for their valuable cooperation and unwavering support throughout the completion of this project.

