## Supplementary material for "A Holistic Approach to Cognitive Frailty in Community-dwelling Older Adults in India: Evidence from a Nationally Representative Survey": . We developed a 20-item frailty index based on physical deficits (Additional File 1),

**Additional 1. The Modified Community-Oriented Frailty Index (COM-FI) for older adults in India**

| **Variable** | **Variable description** | **Measure** | **Coding** | |
| --- | --- | --- | --- | --- |
| **Exhaustion** | Did you feel tired out or low on energy all the time | General health | Yes=1  No=0 | |
| **Low body weight** | Body Mass Index (BMI) in the lowest quintile, adjusted for men and women | General health | Men  BMI≤18.1= 1  BMI>18.1= 0 | Women  BMI≤18.5= 1  BMI>18.5= 0 |
| **Low grip strength** | Handgrip strength in the dominant hand in the lowest quintile for men and women | General health | Men  handgrip ≤18.3= 1  handgrip >18.3= 0 | Women  handgrip ≤11.8= 1  handgrip >11.8= 0 |
| **Low walking speed** | Walking speed in the lowest quintile for men and women | General health | Men  Walk ≥ 4.3min.= 1  Walk < 4.3min.= 0 | Women  Walk ≥ 4.8min.= 1  Walk < 4.8min.= 0 |
| **Low physical activity** | How often do you take part in sports or vigorous activities, such as running or jogging, swimming, going to a health center or gym, cycling, or digging with a spade or shovel, heavy lifting, chopping, farm work, fast bicycling, cycling with loads | General health | One to three times a month / Hardly ever or never= 1  Every day/More than once a week/Once a week=0 | |
| **Hypertension** | Hypertension or high blood pressure (self-reported) | Wellbeing assessment | Yes=1  No=0 | |
| **Diabetes** | Diabetes or high blood sugar (self-reported) | Wellbeing assessment | Yes=1  No=0 | |
| **Lung disease** | Chronic lung disease such as asthma, chronic obstructive pulmonary disease/Chronic bronchitis or other chronic lung problems (self-reported) | Wellbeing assessment | Yes=1  No=0 | |
| **Heart disease** | Chronic heart diseases such as coronary heart disease (heart attack or Myocardial Infarction), congestive heart failure, or other chronic heart problems (self-reported) | Wellbeing assessment | Yes=1  No=0 | |
| **Stroke** | Stroke (self-reported) | Wellbeing assessment | Yes=1  No=0 | |
| **Bone problems** | Arthritis or rheumatism, Osteoporosis or other bone/joint diseases (self-reported) | Wellbeing assessment | Yes=1  No=0 | |
| **Neurological problems** | Any neurological, or psychiatric problems such as depression, unipolar/bipolar disorders, convulsions, Parkinson’s etc. (but excluding dementia) (self-reported) | Wellbeing assessment | Yes=1  No=0 | |
| **Polypharmacy** | How many prescribed medications do you take? | Wellbeing assessment | ≥4 =1  <4 =0 | |
| **Injury** | In the past two years, have you sustained any major injury | Wellbeing assessment | Yes=1  No=0 | |
| **Dressing** | Difficulties dressing, including putting on chappals, shoes, etc. | ADL | Yes=1  No=0 | |
| **Walking** | Difficulties in walking across a room | ADL | Yes=1  No=0 | |
| **Bathing** | Difficulties in bathing | ADL | Yes=1  No=0 | |
| **Eating** | Difficulties in eating | ADL | Yes=1  No=0 | |
| **Bed** | Difficulties in getting in and out of bed | ADL | Yes=1  No=0 | |
| **Toilet use** | Difficulties in using the toilet, including getting up and down | ADL | Yes=1  No=0 | |
